# Single-cell lung eQTL colocalization implicates alveolar epithelial regulatory effects at COPD lung-function loci

**DOI:** 10.64898/2026.11.12.26367877

**Authors:** Rahel Barbara Eberle, Heena Rijhwani, Courtney Tern, Michael H. Cho, Aabida Saferali

## Abstract

**Background:** Chronic obstructive pulmonary disease (COPD) and lung function are highly heritable, but for most genome-wide association study (GWAS) loci, the effector gene and the lung cell type in which it acts are unknown. Single-cell expression quantitative trait loci (eQTLs) offer a way to resolve the cell types in which regulatory variants act.

**Results:** We integrated a multi-ancestry GWAS of the ratio of forced expiratory volume in one second to forced vital capacity (FEV1/FVC; Shrine et al., n = 588,452) with single-cell lung eQTLs across 38 cell types (Natri et al., n = 114) by colocalization. We identified 20 candidate colocalizations (posterior probability of a shared causal variant > 0.7) for 13 genes in nine cell types, predominantly epithelial and immune. The two strongest signals localized DSP, an established COPD effector gene, and NRP2, a candidate effector gene, to alveolar type 2 epithelial cells; only these two colocalizations remained supported under a more conservative shared-effect prior. Both signals, together with FAM13A, were also detectable in bulk lung eQTLs, whereas most remaining loci colocalized only in the single-cell analysis.

**Conclusions:** Single-cell eQTL colocalization resolves the alveolar epithelium as a cellular context for DSP and nominates NRP2 as a candidate lung function effector gene in the same compartment, highlighting the additional cellular resolution provided by single-cell data. The remaining candidate colocalizations warrant replication.

## Background

Chronic obstructive pulmonary disease (COPD) is a common respiratory disease with a substantial genetic component, arising from gene by environment interactions [1].

Genome-wide association studies (GWAS) of COPD and of the ratio of forced expiratory volume in one second to forced vital capacity (FEV1/FVC), the lung function trait most relevant to COPD, have identified many susceptibility loci and overlap substantially with one another [2, 3, 4]. Translating these loci into disease mechanisms requires identifying both the effector gene at each locus and the lung cell type in which its regulatory effect occurs. Integrative genetic studies have begun to nominate effector genes, such as DSP [4], but because bulk tissue averages expression across many cell types, bulk approaches cannot directly resolve the cell type in which a regulatory variant acts. The recent availability of large single-cell lung eQTL resources now makes this resolution possible [5]. Here, we integrate a multi-ancestry FEV1/FVC GWAS with single-cell lung eQTLs across 38 cell types to identify candidate effector genes for lung function and the lung cell populations in which they act.

## Methods

We obtained single-cell lung eQTL summary statistics from Natri et al. [5], comprising cis-eQTLs from pseudobulk single-cell RNA sequencing of lung tissue from 114 individuals (66 with pulmonary fibrosis, 48 unaffected) across 38 cell types, of mixed ancestry. Lung function summary statistics were from the multi-ancestry FEV1/FVC meta-analysis of Shrine et al. [3] (588,452 participants; 399 genome-wide significant FEV1/FVC signals; see Additional file 1: Supplementary Methods). For comparison with bulk expression, we used published lung eQTL data from 1,291 Lung Tissue Research Consortium (LTRC) samples generated within TOPMed [6].

We defined genomic regions by extending 100 kb on either side of each genome-wide significant GWAS variant, and restricted each region to variants present in both datasets. Each genome-wide significant FEV1/FVC signal defined its own region, centered on that variant and extended 100 kb on either side; where signals were close enough that their windows overlapped, each was tested independently on its own centered window rather than collapsed to a single representative. This window focuses the analysis on local association signals while limiting neighboring independent signals, given the single-causal-variant assumption of coloc; a window-size sensitivity analysis for the DSP and NRP2 loci is reported in Additional file 1. We tested each region for colocalization between the GWAS and the single-cell eQTL of each gene in each cell type (workflow in Additional file 1: Figure S1) using the coloc R package [7], which estimates posterior probabilities for five hypotheses; hypothesis 4 (PPH4) corresponds to a shared causal variant. We used prior probabilities of 1×10^−4^ for association with each trait and 1×10^−5^ for a shared effect, and defined evidence for colocalization as PPH4 > 0.7, consistent with thresholds used in previous studies [8]. Colocalizations in regions with fewer than 100 variants were considered preliminary. We assessed robustness across shared-effect priors, and repeated the analysis with the bulk LTRC eQTL data. Analyses used R version 4.1.2 [9] and the coloc R package version 5.2.3 [7].

## Results

Across 142 genome-wide significant regions containing 24,523 variants and 1,280 genes, we tested colocalization between the FEV1/FVC GWAS and single-cell eQTLs in each of 38 lung cell types. Under our primary model, we identified 20 candidate region, gene, and cell type colocalizations at PPH4 > 0.7, representing 15 genomic regions (Table 1). Colocalizations were concentrated in epithelial and immune cell types, and the two strongest converged on alveolar epithelial cells.

**Table 1.** Candidate colocalizations of FEV1/FVC with single-cell lung eQTLs and corresponding bulk results.

| **Gene** | **Cell type** | **Region SNP** | **Single-cell PPH4** | **Bulk lung PPH4** |
|---|---|---|---|---|
| DSP | epithelial AT2 | rs2076295 | >0.99 | 1.00 |
| NRP2 | epithelial AT2 | rs2241153 | >0.99 | 0.9995 |
| DYDC2 | epithelial Ciliated | rs10748526 | 0.94 | 0.65 |
| ANXA3 | epithelial Secretory-SCGB1A1+MUC5B+ | rs4975128 | 0.93 | 0.13 |
| ATP5MK | epithelial Ciliated | rs11594179 | 0.93* | 0.16 |
| DSP | epithelial AT1 | rs2076295 | 0.89 | 1.00 |
| DYDC2 | epithelial DifferentiatingCiliated | rs10748526 | 0.89 | 0.65 |
| DRAM2 | epithelial Ciliated | rs10857859 / rs629619 | 0.88 | <0.001 |
| TNRC6B | mesenchymal SMC | rs77516118 | 0.87 | 0.28 |
| NRP2 | epithelial AT1 | rs2241153 | 0.82 | 0.9995 |
| KAT7 | immune Interstitial macrophage | rs7221118 | 0.80* | 0.05 |
| WDR77 | immune Inflammatory monocyte | rs10857859 / rs629619 | 0.80 | 0.04 |
| RHOV | epithelial DifferentiatingCiliated | rs9944249 | 0.78* | 0.17 |
| DRAM2 | immune Monocyte-derived macrophage | rs10857859 / rs629619 | 0.72 | <0.001 |
| LDLRAP1 | epithelial AT1 | rs9438876 | 0.72 | 0.05 |
| RAMP1 | endothelial Systemic venous | rs11677932 | 0.72 | 0.05 |
| FAM13A | epithelial AT2 | rs1246642 | 0.71 | 0.905 |

Candidate colocalizations are those with single-cell PPH4 > 0.7; rows are ordered by single-cell PPH4. \* marks regions with fewer than 100 tested variants. For DRAM2 and WDR77, two region-defining SNPs 4384 bp apart gave near-identical results and are shown as one row per cell type, so the 20 region, gene, and cell type colocalizations appear as 17 rows. Bulk lung PPH4 is the colocalization of the same region and gene using LTRC/TOPMed bulk lung eQTLs. Effect alleles and directions of effect are given in Additional file 1: Table S1.

The strongest colocalization localized DSP, a previously reported COPD and lung function effector gene, to alveolar epithelial cells. FEV1/FVC and DSP expression colocalized in alveolar type 2 (AT2) cells with PPH4 exceeding 0.99, with the same signal in alveolar type 1 (AT1) cells (rs2076295, PPH4 0.89; Figure 1). The rs2076295 T allele was associated with lower FEV1/FVC and higher DSP expression.

**Figure 1.**
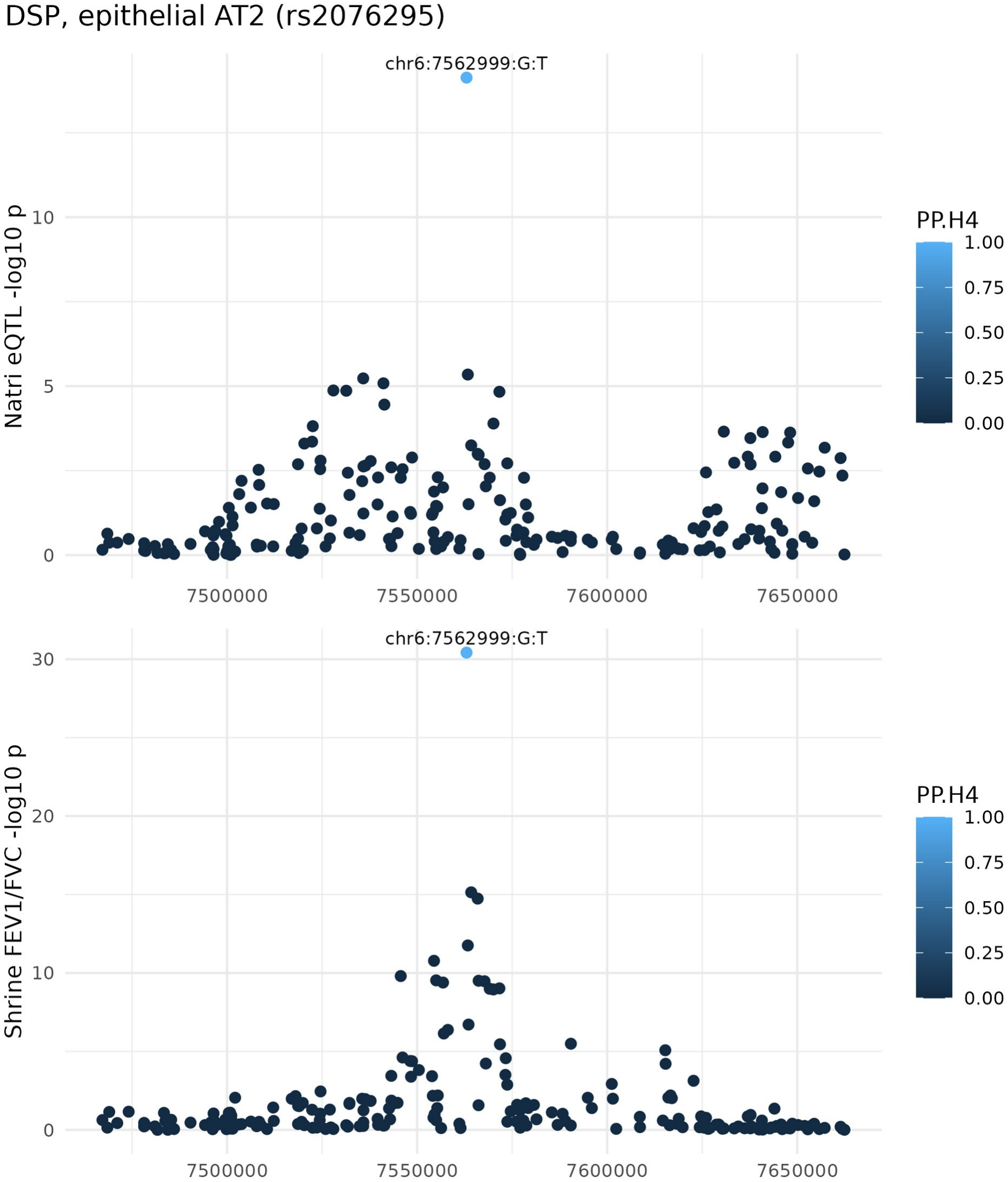
Colocalization of DSP expression in alveolar type 2 cells with FEV1/FVC. Association of variants within 100 kb of rs2076295 with DSP expression in epithelial AT2 cells (single-cell eQTL, Natri et al.; upper panel) and with FEV1/FVC (GWAS, Shrine et al.; lower panel). Each point is one variant, plotted by GRCh38 position on chromosome 6 and -log10 p-value, and colored by its per-SNP posterior probability of being the shared causal variant (PP.H4). The labeled variant, chr6:7562999:G:T (rs2076295), has the highest per-SNP posterior probability.

The strongest colocalization at a locus not previously established as a lung function effector gene localized NRP2 to alveolar epithelial cells, in AT2 cells with PPH4 exceeding 0.99 and with weaker support in AT1 cells (rs2241153, PPH4 0.82; Figure 2). The rs2241153 C allele was associated with higher FEV1/FVC and higher NRP2 expression.

**Figure 2.**
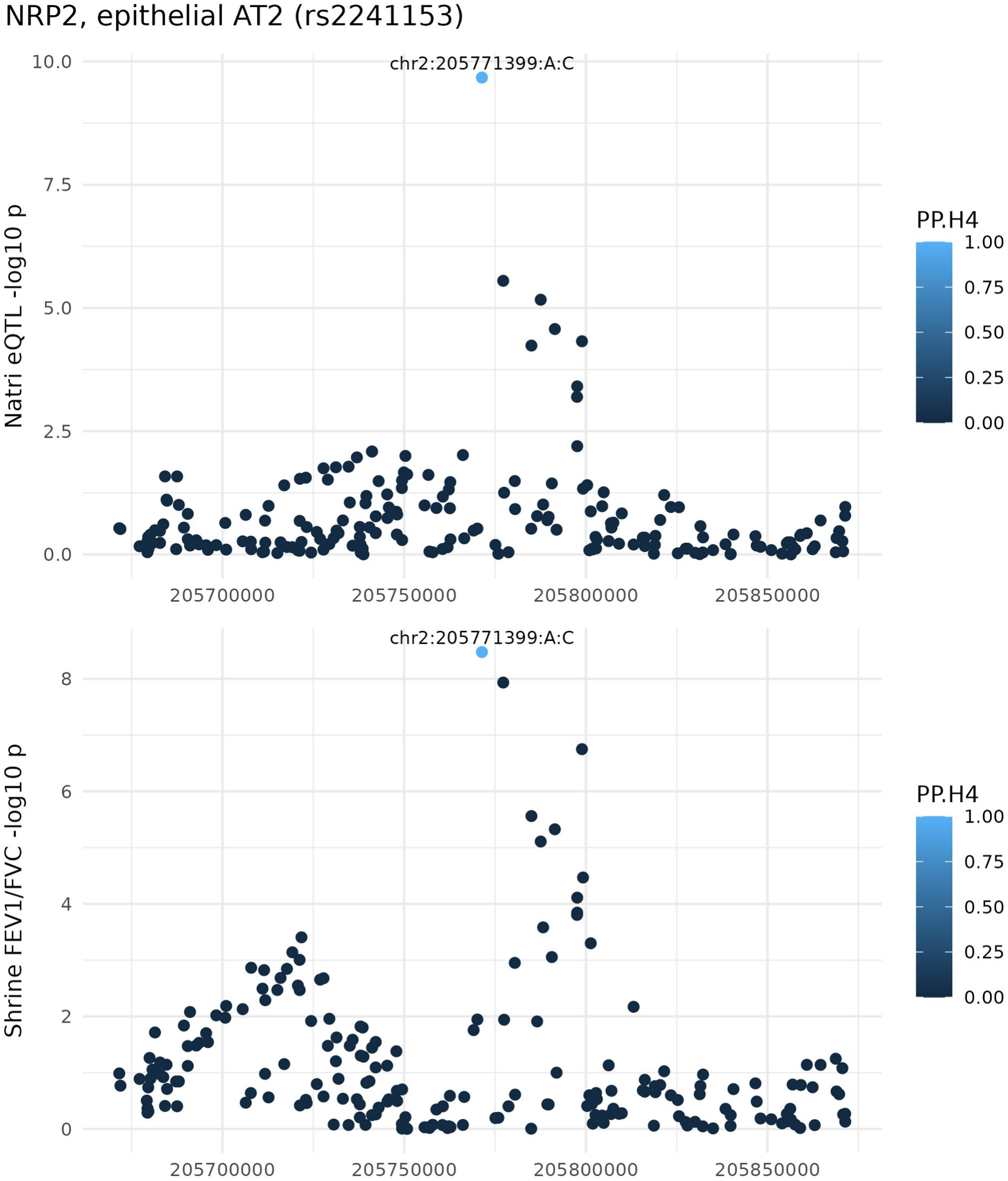
Colocalization of NRP2 expression in alveolar type 2 cells with FEV1/FVC. Association of variants within 100 kb of rs2241153 with NRP2 expression in epithelial AT2 cells (single-cell eQTL, Natri et al.; upper panel) and with FEV1/FVC (GWAS, Shrine et al.; lower panel). Each point is one variant, plotted by GRCh38 position on chromosome 2 and -log10 p-value, and colored by its per-SNP posterior probability of being the shared causal variant (PP.H4). The labeled variant, chr2:205771399:A:C (rs2241153), has the highest per-SNP posterior probability.

The remaining colocalizations, in additional epithelial, immune, mesenchymal, and endothelial cell types, are listed in Table 1 and described in Additional file 1 (Table S1, Figures S2-S4); DSP and NRP2 were identified through an unbiased genome-wide scan rather than by candidate-gene selection. Three of the 15 colocalizing regions were also recovered using bulk lung eQTL data, namely DSP, NRP2, and FAM13A, an established COPD gene [10] (Table 1; Additional file 1: Table S2). For the remaining regions, the colocalization was detected only in the single-cell analysis, highlighting the additional cellular resolution provided by the single-cell data. For example, we found PPH4 > 0.9 for DYDC2 and ATP5MK, both in epithelial ciliated cells, as well as ANXA3 in epithelial secretory cells; these all had much weaker evidence of colocalization in bulk data (PPH4 = 0.13-0.65).

The colocalizations were sensitive to analytic choices in an informative way. Because the shared-effect prior is the most influential and least-calibrated coloc parameter [11], we assessed sensitivity to it; at a more conservative prior (1×10^−6^), only the DSP (AT2) and NRP2 (AT2) colocalizations retained PPH4 > 0.7 (Additional file 1: Figure S5). Both were robust to a 250-kb window and the NRP2 (AT2) result to a 500-kb window; at 500 kb the DSP locus admitted a second independent association, inconsistent with the single-causal-variant assumption of coloc (Additional file 1: Table S4). We therefore regard the AT2 colocalizations for DSP and NRP2 as our most robust findings, and the remaining colocalizations as candidates that warrant replication.

## Discussion

Integrating a multi-ancestry lung function GWAS with single-cell lung eQTLs across 38 cell types, we identified cell-type-specific regulatory associations at COPD-relevant lung function loci. The strongest and most prior-robust signals converged on alveolar epithelial cells and involved two genes, the established effector gene DSP and the candidate gene NRP2. Recovering DSP, a known lung function effector gene, and resolving its regulatory effect to the alveolar epithelium supports the validity of the approach; nominating NRP2 in the same compartment illustrates how single-cell regulatory data can move a GWAS locus toward a specific gene and cell type.

DSP encodes desmoplakin, which regulates cell-cell junctions and the cigarette-smoke injury response in alveolar type 2 epithelial cells [12], and the rs2076295 region regulates DSP expression in airway epithelium [13]. rs2076295 is associated with COPD and idiopathic pulmonary fibrosis in opposite directions, the major T allele increasing DSP expression and COPD risk [4, 13, 14, 15]; consistently, the T allele was associated with higher DSP expression and lower FEV1/FVC in our data. Prior work established DSP as an effector gene at this locus; our single-cell analysis adds that this regulatory effect is detectable in alveolar type 1 and type 2 epithelial cells, identifying the alveolar epithelium as a cellular context for the DSP effect on lung function.

NRP2 encodes neuropilin 2, a co-receptor for class 3 semaphorin and vascular endothelial growth factor ligands. In the lung, neuropilin 2 is expressed in respiratory epithelium and participates in semaphorin-mediated branching morphogenesis during development [16], and also regulates airway inflammatory responses [17]. To our knowledge, NRP2 has not previously been established as an effector gene for COPD or lung function. Our colocalization nominates NRP2 as a candidate effector gene acting in alveolar epithelial cells; although its expression in lung epithelium is consistent with this assignment, a specific role in adult alveolar epithelial regulation of lung function will require functional validation. Together, DSP and NRP2 illustrate complementary uses of single-cell eQTL colocalization: resolving the cellular context of an established effector gene, and nominating a candidate effector gene in a disease-relevant cell type.

Several considerations bound these results. The coloc method assumes a single causal variant per region, which can miss loci with multiple independent signals; methods such as coloc-SuSiE relax this assumption [18]. The GWAS and single-cell eQTL datasets are of mixed and non-identical ancestry, which may reduce colocalization power [19], and differences between the single-cell and bulk results may reflect sample size, genotype density, composition, and eQTL architecture in addition to cellular resolution. Our colocalizations are also sensitive to the shared-effect prior, the least-certain coloc parameter: only DSP (AT2) and NRP2 (AT2) survived a conservative prior, so we present these as most robust and the remainder as prior-dependent candidates rather than likely false positives. Finally, the analysis is statistical and does not establish causal gene function; replication in independent single-cell eQTL data and functional validation are needed.

## Conclusions

Colocalization of a multi-ancestry FEV1/FVC GWAS with single-cell lung eQTLs identified 20 candidate cell-type-specific regulatory effects at lung function loci, the most robust of which localized DSP and NRP2 to alveolar type 2 epithelial cells. Cell-resolved regulatory genetics can help move COPD lung function GWAS loci from variant to gene to relevant lung cell population, here most robustly implicating DSP and NRP2 in alveolar epithelial cells. Replication in independent single-cell eQTL data and functional validation are needed to establish causal gene function.

## Data Availability

All data analyzed in this study are from previously published studies. The multi-ancestry FEV1/FVC GWAS summary statistics of Shrine et al. are available from the GWAS Catalog under accession GCST90244094. The single-cell lung eQTL summary statistics of Natri et al. are available from the Gene Expression Omnibus under accession GSE227136. The bulk LTRC/TOPMed lung eQTL data are described in Orchard et al. (Science 2026;393:eadx2989). Analysis code is available from the corresponding author on reasonable request.

## Abbreviations

AT1: alveolar type 1
AT2: alveolar type 2
COPD: chronic obstructive pulmonary disease
eQTL: expression quantitative trait locus
FEV1/FVC: ratio of forced expiratory volume in one second to forced vital capacity
GWAS: genome-wide association study
LTRC: Lung Tissue Research Consortium
PPH4: posterior probability of hypothesis 4 (a shared causal variant)
SNP: single-nucleotide polymorphism
TOPMed: Trans-Omics for Precision Medicine

## Declarations

### Ethics approval and consent to participate

This study analyzed summary-level data only; no individual-level human data were generated or accessed for this work. The GWAS and single-cell eQTL summary statistics were publicly available from previously published studies [3, 5], each of which obtained ethical approval and informed consent from participants as described in the original publications. Bulk lung eQTL summary statistics were derived from the Lung Tissue Research Consortium (LTRC) within the NHLBI TOPMed program [6]. Analysis of LTRC data at Brigham and Women’s Hospital was approved by the Mass General Brigham Institutional Review Board (protocol 2018P000186), and all LTRC participants provided written informed consent.

### Consent for publication

Not applicable.

### Availability of data and materials

All data analyzed in this study are from previously published studies. The multi-ancestry FEV1/FVC GWAS summary statistics of Shrine et al. [3] are available from the GWAS Catalog under accession GCST90244094 [20]. The single-cell lung eQTL summary statistics of Natri et al. [5] are available from the Gene Expression Omnibus under accession GSE227136 [21]. The bulk LTRC/TOPMed lung eQTL data are described in [6]. Analysis code is available from the corresponding author on reasonable request.

### Competing interests

MHC reports grant funding from Bayer and consulting fees from Apogee Therapeutics and BMS, unrelated to the current work. RBE is currently employed by Genetica AG, unrelated to this work. The other authors declare that they have no competing interests.

### Funding

AS was supported by NHLBI K01HL157613 and R03HL183107. MHC was supported by NHLBI R01HL168199, R01HL162813, R01HL153248, and R01HL135142. RBE was supported by the Swiss Lung Foundation. The funders had no role in the design of the study, the analysis and interpretation of data, or the writing of the manuscript.

### Authors’ contributions

RBE and HR performed the analyses. AS and MHC conceived and supervised the study. CT contributed to data processing. RBE and AS wrote the manuscript. All authors read and approved the final manuscript.

## Acknowledgements

This work is based on the master thesis of RBE.

## Additional files

**Additional file 1**. File name: Additional_file_1.docx. File format: Microsoft Word (.docx). Title: Supplementary Materials. Description: Supplementary Methods (summary statistics, data harmonization, region definition, region-window sensitivity); Supplementary Results (additional colocalizations); Tables S1-S4; Figures S1-S5; Supplementary References.

## Notes

### Author Declarations

The Institutional Review Board of Mass General Brigham (Massachusetts, USA) gave ethical approval for this work (protocol 2018P000186, Network Analysis of Multiple Omics Data in the Lung Tissue Research Consortium). This study analyzed summary-level data only. The GWAS and single-cell eQTL summary statistics were openly available before the initiation of the study (GWAS Catalog GCST90244094; Gene Expression Omnibus GSE227136). Bulk lung eQTL summary statistics were derived from the Lung Tissue Research Consortium within the NHLBI Trans-Omics for Precision Medicine (TOPMed) program.

