## Supplement for "Single-cell lung eQTL colocalization implicates alveolar epithelial regulatory effects at COPD lung-function loci"

### Additional file 1: Supplementary Materials

*Supplementary Materials. Tables S1-S4 and Figures S1-S5 support the main text.*

#### Supplementary Methods

**Summary statistics.** We used the released summary statistics of Shrine et al on untransformed FEV1/FVC. Published results included additional signals based on rank-inverse-normal transformation, which produced minor discrepancies in p-values and effect estimates. The p-values from the transformed and untransformed data were highly correlated (Spearman correlation 0.88), and 55 signals reported as genome-wide significant in the paper had p-values above the study's genome-wide significance threshold in the released dataset. Because the discrepancies are minor and the released untransformed statistics are the publicly available data, we therefore retained the released summary statistics for all variants in the colocalization analysis.

Natri eQTL summary statistics were filtered to minor allele frequency above 5%, Hardy-Weinberg equilibrium p-value above $10^{-6}$, and pruned for high correlation, yielding approximately 1.9 million variants.

**Data harmonization:** Both datasets were analyzed on genome build GRCh38: the Natri single-cell eQTL summary statistics were reported on GRCh38, and the publicly available Shrine GWAS summary statistics, released on GRCh37, were lifted over to GRCh38 prior to analysis. We matched variants between the single-cell eQTL and GWAS data by chromosome and position. Natri et al. reported only the assessed allele, without an explicit other or reference allele. For each matched variant we compared the Natri assessed allele with the Shrine effect and other alleles: where the assessed allele matched the Shrine other allele, we swapped the Shrine effect and other alleles and reversed the sign of the Shrine effect estimate, so that all effect estimates are oriented to the Natri assessed allele. Variants whose assessed allele matched neither Shrine allele, or matched both, were removed, and minor allele frequency was computed from the reported effect allele frequency assuming biallelic single-nucleotide polymorphisms (SNPs).

**Region definition.** We tested the 399 genome-wide significant FEV1/FVC signals reported by Shrine et al. in the mixed-ancestry analysis. Each signal defined its own region, centered on that variant and extended 100 kb on either side, and each region was restricted to variants present in both the GWAS and the single-cell eQTL data. Of the 399 signals, 142 had their region-defining variant present in the single-cell eQTL data in at least one cell type and were carried into the colocalization analysis; the remaining 257 were dropped because no shared variant was available.

**Region-window sensitivity.** The primary analysis defined each region as ±100 kb around the lead GWAS variant. To assess sensitivity to this choice, we repeated the DSP and NRP2 colocalizations in AT2 and AT1 cells using ±250 kb and ±500 kb windows, with all other parameters unchanged (Table S4). The recomputed ±100 kb values reproduced the primary results exactly. The NRP2 (AT2) colocalization was essentially unchanged across all three windows (PPH4 = 0.999), and the DSP (AT2) colocalization remained very strong at ±100 kb and ±250 kb (PPH4 = 1.00). At ±500 kb the DSP colocalization weakened (PPH4 = 0.63 in AT2 and 0.09 in AT1): the wider window admitted a second, independent genome-wide significant lung function association at this locus (lead variant rs11759956), so under the single-causal-variant assumption of coloc probability shifts toward distinct causal variants (PP.H3 = 0.37 in AT2), even though rs2076295 remained the variant with the highest per-SNP posterior probability of a shared effect. This behavior illustrates the importance of defining regions to isolate individual association signals when applying a single-causal-variant colocalization model, and indicates that single-variant colocalization becomes less appropriate as the window widens at loci carrying multiple independent signals. Thus, the DSP and NRP2 conclusions were unchanged at the primary window and robust to a ±250 kb window; the NRP2 (AT2) result was additionally robust to a ±500 kb window.

#### Supplementary Results: additional colocalizations

Beyond DSP and NRP2, the primary model identified additional candidate colocalizations, listed in full with their cell types, variants, PPH4 values, and effect directions in Table S1 (colocalization plots in Supplementary Figure S4). These are reported for completeness and require replication; we note only directly relevant prior evidence for a subset.

**FAM13A** colocalized in epithelial AT2 cells (rs1246642) and, together with DSP and NRP2, was recovered in the bulk analysis; it is an established COPD gene with a characterized role in AT2 cells [S1, S2].

**DYDC2** colocalized in epithelial ciliated and differentiating ciliated cells (rs10748526; top variant rs12220642); it is expressed in bronchial ciliated cells and has previously been nominated as a candidate causal gene for COPD from integrated GWAS and eQTL data [S3, S4].

**DRAM2** colocalized in epithelial ciliated and monocyte-derived macrophage cells (rs10857859), a variant previously reported as a monocyte eQTL for DRAM2 [S5].

**KAT7** colocalized in immune interstitial macrophages (rs7221118) and has been reported as downregulated in COPD airway epithelium [S6].

The remaining candidate colocalizations (ANXA3, ATP5MK, TNRC6B, WDR77, RHOV, LDLRAP1, and RAMP1) are reported in Table S1.

#### Supplementary Tables

**Table S1.** All 20 region, gene, and cell type colocalizations at PPH4 > 0.7. * marks regions with fewer than 100 tested variants. FEV1/FVC and expression directions are for the effect allele at the SNP with the highest per-SNP posterior probability of a shared causal variant. Genes in the rs10857859 and rs629619 rows share one locus (the two region-defining SNPs are 4,384 bp apart and give near-identical results).

| Gene | Cell type | Region SNP | Top-PP SNP | PPH4 | SNPs | Effect allele | FEV1/FVC | Expression |
|---|---|---|---|---|---|---|---|---|
| DSP | epithelial AT2 | rs2076295 | rs2076295 | >0.99 | 210 | T | lower | higher |
| NRP2 | epithelial AT2 | rs2241153 | rs2241153 | >0.99 | 199 | C | higher | higher |
| DYDC2 | epithelial Ciliated | rs10748526 | rs12220642 | 0.94 | 157 | T | lower | higher |
| ANXA3 | epithelial Secretory-SCGB1A1+MUC5B+ | rs4975128 | rs4975128 | 0.93 | 114 | G | higher | lower |
| ATP5MK | epithelial Ciliated | rs11594179 | rs11594179 | 0.93* | 57 | C | higher | higher |
| DSP | epithelial AT1 | rs2076295 | rs2076295 | 0.89 | 204 | T | lower | higher |
| DYDC2 | epithelial DifferentiatingCiliated | rs10748526 | rs12220642 | 0.89 | 156 | T | lower | higher |
| DRAM2 | epithelial Ciliated | rs10857859 | rs10857859 | 0.88 | 168 | G | lower | lower |
| DRAM2 | epithelial Ciliated | rs629619 | rs10857859 | 0.88 | 167 | G | lower | lower |
| TNRC6B | mesenchymal SMC | rs77516118 | rs77516118 | 0.87 | 193 | T | higher | higher |
| NRP2 | epithelial AT1 | rs2241153 | rs2241153 | 0.82 | 195 | C | higher | higher |
| KAT7 | immune Interstitial macrophage | rs7221118 | rs7221118 | 0.80* | 95 | T | lower | lower |
| WDR77 | immune Inflammatory monocyte | rs10857859 | rs10857859 | 0.80 | 166 | G | lower | lower |
| WDR77 | immune Inflammatory monocyte | rs629619 | rs10857859 | 0.80 | 165 | G | lower | lower |
| RHOV | epithelial DifferentiatingCiliated | rs9944249 | rs9944249 | 0.78* | 98 | T | higher | higher |
| DRAM2 | immune Monocyte-derived macrophage | rs10857859 | rs10857859 | 0.72 | 168 | G | lower | lower |
| DRAM2 | immune Monocyte-derived macrophage | rs629619 | rs10857859 | 0.72 | 167 | G | lower | lower |

| | | | | | | | | |
|---|---|---|---|---|---|---|---|---|
| LDLRAP1 | epithelial AT1 | rs9438876 | rs9438876 | 0.72 | 187 | A | higher | lower |
| RAMP1 | endothelial Systemic venous | rs11677932 | rs11677932 | 0.72 | 138 | G | lower | lower |
| FAM13A | epithelial AT2 | rs1246642 | rs1246642 | 0.71 | 101 | C | higher | lower |

**Table S2.** Colocalization of FEV1/FVC with bulk LTRC/TOPMed lung eQTL data for the 15 region, gene pairs significant in the single-cell analysis. Only three region-gene pairs also reached PPH4 > 0.7 using bulk lung eQTLs.

| Gene | Region SNP | Bulk PPH4 |
|---|---|---|
| DSP | rs2076295 | 1.00 |
| NRP2 | rs2241153 | 0.9995 |
| FAM13A | rs1246642 | 0.905 |
| TNRC6B | rs77516118 | 0.28 |
| RHOV | rs9944249 | 0.17 |
| ATP5MK | rs11594179 | 0.16 |
| ANXA3 | rs4975128 | 0.13 |
| DYDC2 | rs10748526 | 0.65 |
| LDLRAP1 | rs9438876 | 0.05 |
| RAMP1 | rs11677932 | 0.05 |
| KAT7 | rs7221118 | 0.05 |
| WDR77 | rs10857859 / rs629619 | 0.04 |
| DRAM2 | rs10857859 / rs629619 | <0.001 |

**Table S3.** Per-cell-type descriptive statistics across all tested regions: number of tested regions, median, minimum, and maximum variants per region, and the distribution of PPH4.

| Cell type | Tested regions | Median SNPs per region | Min SNPs | Max SNPs | Genes | Median PPH4 | Max PPH4 |
|---|---|---|---|---|---|---|---|
| endothelial aCap | 137 | 131 | 11 | 1579 | 1237 | 0.08 | 0.59 |
| endothelial arteriole | 138 | 130 | 12 | 1545 | 1253 | 0.08 | 0.58 |
| endothelial gCap | 128 | 130 | 11 | 1545 | 891 | 0.08 | 0.23 |
| endothelial Lymphatic | 141 | 131 | 10 | 1551 | 1275 | 0.08 | 0.65 |
| endothelial Systemicvenous | 134 | 130 | 13 | 1496 | 1217 | 0.08 | 0.72 |
| endothelial venule | 138 | 135 | 13 | 1544 | 1254 | 0.08 | 0.53 |
| epithelial AT1 | 133 | 130 | 13 | 1560 | 1217 | 0.08 | 0.89 |
| epithelial AT2 | 137 | 136 | 14 | 1561 | 1249 | 0.08 | >0.99 |
| epithelial Basal | 132 | 127 | 13 | 1452 | 1221 | 0.08 | 0.44 |
| epithelial Ciliated | 138 | 134 | 13 | 1503 | 1251 | 0.08 | 0.94 |
| epithelial DifferentiatingCiliated | 134 | 135 | 12 | 1547 | 1172 | 0.08 | 0.89 |
| epithelial KRT5-KRT17+ | 130 | 123 | 12 | 1480 | 933 | 0.08 | 0.31 |
| epithelial | 136 | 132 | 11 | 1537 | 1238 | 0.08 | 0.34 |

| | | | | | | | |
|---|---|---|---|---|---|---|---|
| Proliferating | | | | | | | |
| epithelial Secretory-SCGB1A1+MUC5B+ | 135 | 133.5 | 12 | 1516 | 1235 | 0.08 | 0.93 |
| epithelial Secretory-SCGB1A1+SCGB3A2+ | 132 | 133 | 13 | 1505 | 1199 | 0.08 | 0.49 |
| epithelial Secretory-SCGB3A2+ | 136 | 135 | 12 | 1507 | 1228 | 0.08 | 0.40 |
| epithelial TransitionalAT2 | 137 | 126 | 11 | 1496 | 1245 | 0.08 | 0.69 |
| immune Alveolarmacrophage | 136 | 133 | 12 | 1522 | 1218 | 0.08 | 0.36 |
| immune Bcells | 130 | 131 | 12 | 1542 | 927 | 0.08 | 0.36 |
| immune CD4 | 135 | 127 | 12 | 1541 | 1242 | 0.08 | 0.31 |
| immune CD8NKT | 136 | 131 | 13 | 1558 | 1234 | 0.08 | 0.46 |
| immune cDC1 | 131 | 124 | 13 | 1476 | 907 | 0.08 | 0.35 |
| immune cDC2 | 138 | 131 | 12 | 1531 | 1251 | 0.08 | 0.54 |
| immune Inflammatory monocyte | 137 | 136 | 13 | 1545 | 1247 | 0.08 | 0.80 |
| immune Interstitialmacrophage | 133 | 126 | 13 | 1515 | 1219 | 0.08 | 0.80 |
| immune Mast | 136 | 128 | 12 | 1517 | 1215 | 0.08 | 0.66 |
| immune moDC | 135 | 137 | 12 | 1557 | 1210 | 0.08 | 0.44 |
| immune Monocyte | 135 | 133 | 13 | 1561 | 1241 | 0.08 | 0.63 |
| immune Monocyte-derivedmacrophage | 139 | 135 | 13 | 1549 | 1260 | 0.08 | 0.72 |
| immune NK | 135 | 128 | 13 | 1536 | 1242 | 0.08 | 0.48 |
| immune pDC | 128 | 123 | 11 | 1420 | 886 | 0.08 | 0.41 |
| immune Plasma | 131 | 127 | 10 | 1497 | 909 | 0.08 | 0.49 |
| immune Proliferating | 136 | 132 | 13 | 1517 | 1246 | 0.08 | 0.63 |
| mesenchymal AdventitialFB | 134 | 131.5 | 12 | 1528 | 1231 | 0.08 | 0.31 |
| mesenchymal AlveolarFB | 130 | 127.5 | 11 | 1498 | 920 | 0.08 | 0.47 |
| mesenchymal Mesothelial | 125 | 120 | 12 | 1452 | 910 | 0.08 | 0.52 |
| mesenchymal Pericyte | 130 | 130 | 13 | 1514 | 921 | 0.08 | 0.27 |
| mesenchymal SMC | 132 | 135 | 12 | 1495 | 935 | 0.08 | 0.87 |
| Total | 142 | 130 | 10 | 1579 | 1280 | 0.08 | >0.99 |

**Table S4.** Region-window sensitivity for the DSP and NRP2 colocalizations (PPH4) at ±100 kb (primary), ±250 kb, and ±500 kb.

| Gene | Cell type | ±100 kb | ±250 kb | ±500 kb |
|---|---|---|---|---|

| | | | | |
|---|---|---|---|---|
| DSP | AT2 | 1.00 | 1.00 | 0.63 |
| DSP | AT1 | 0.89 | 0.90 | 0.09 |
| NRP2 | AT2 | 0.9994 | 0.9994 | 0.9994 |
| NRP2 | AT1 | 0.82 | 0.83 | 0.82 |

#### Supplementary Figures

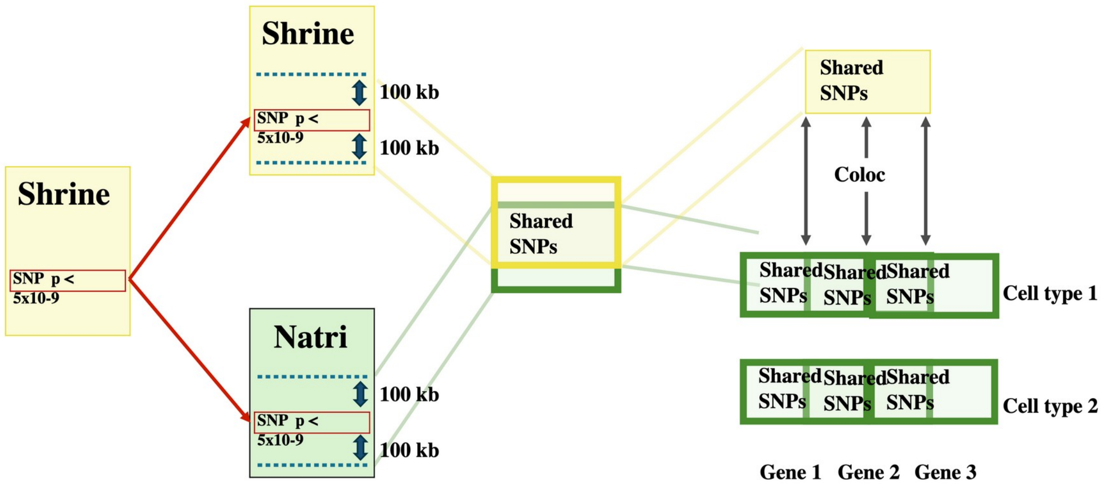


**Figure S1.** Analysis workflow.

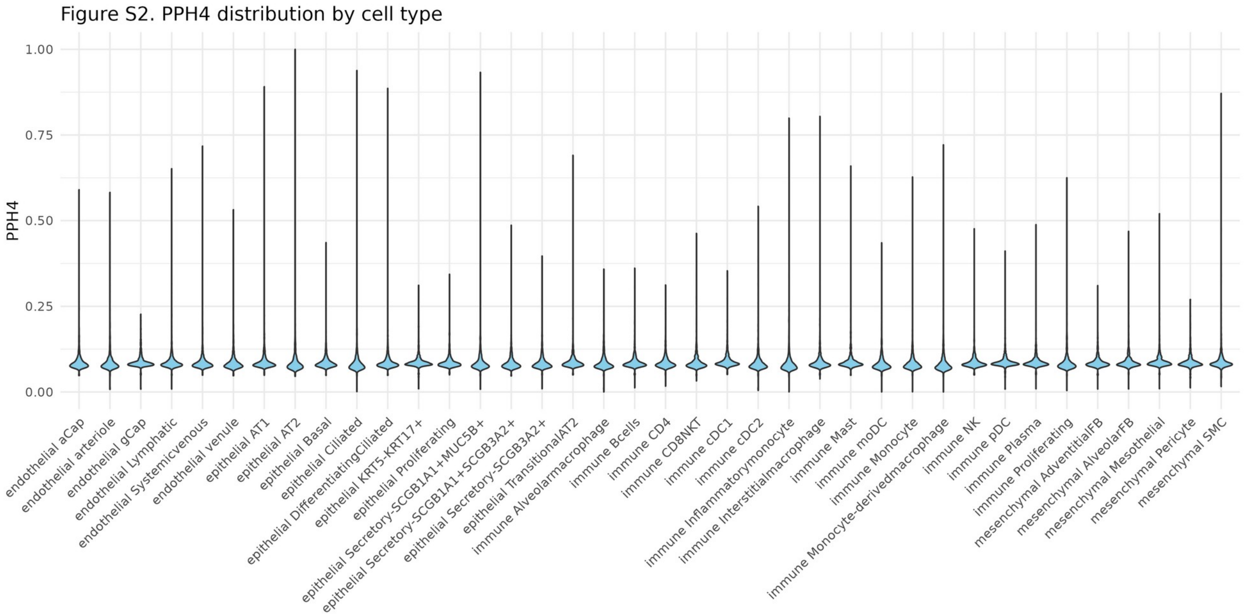


**Figure S2.** Distribution of PPH4 across all tested region, gene, and cell type combinations (violin plot per cell type; median PPH4 0.08 overall).

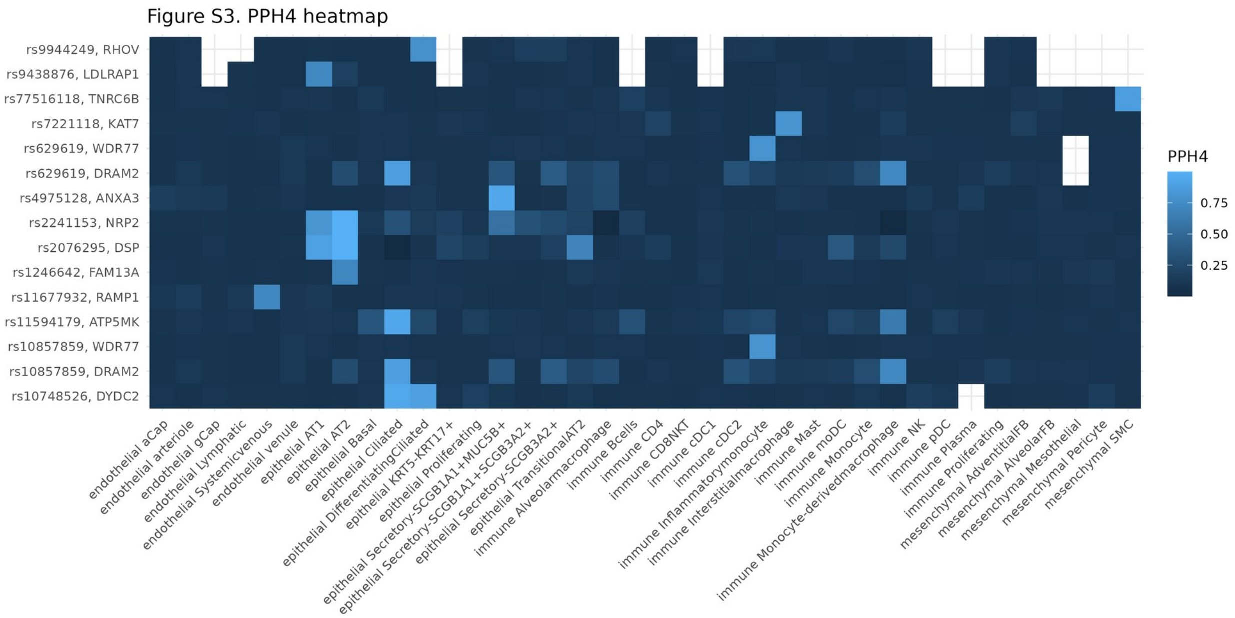


**Figure S3.** PPH4 heatmap across cell types for each region reaching PPH4 > 0.7 in at least one cell type, showing accumulation in epithelial, and to a lesser extent immune, cell types.

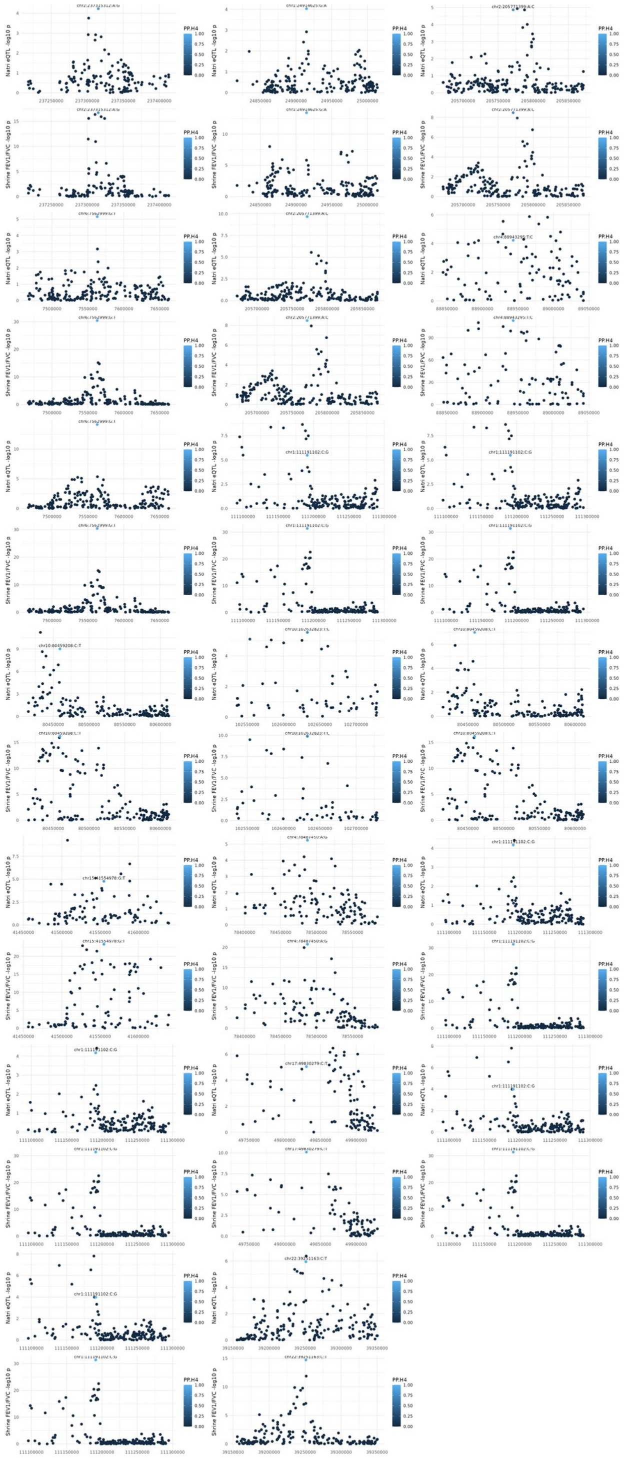

**Figure S4.** Multi-panel colocalization plots for the 20 candidate region, gene, and cell type pairs in Table S1, each showing the single-cell eQTL and FEV1/FVC associations across the region colored by the per-SNP posterior probability of a shared causal variant.

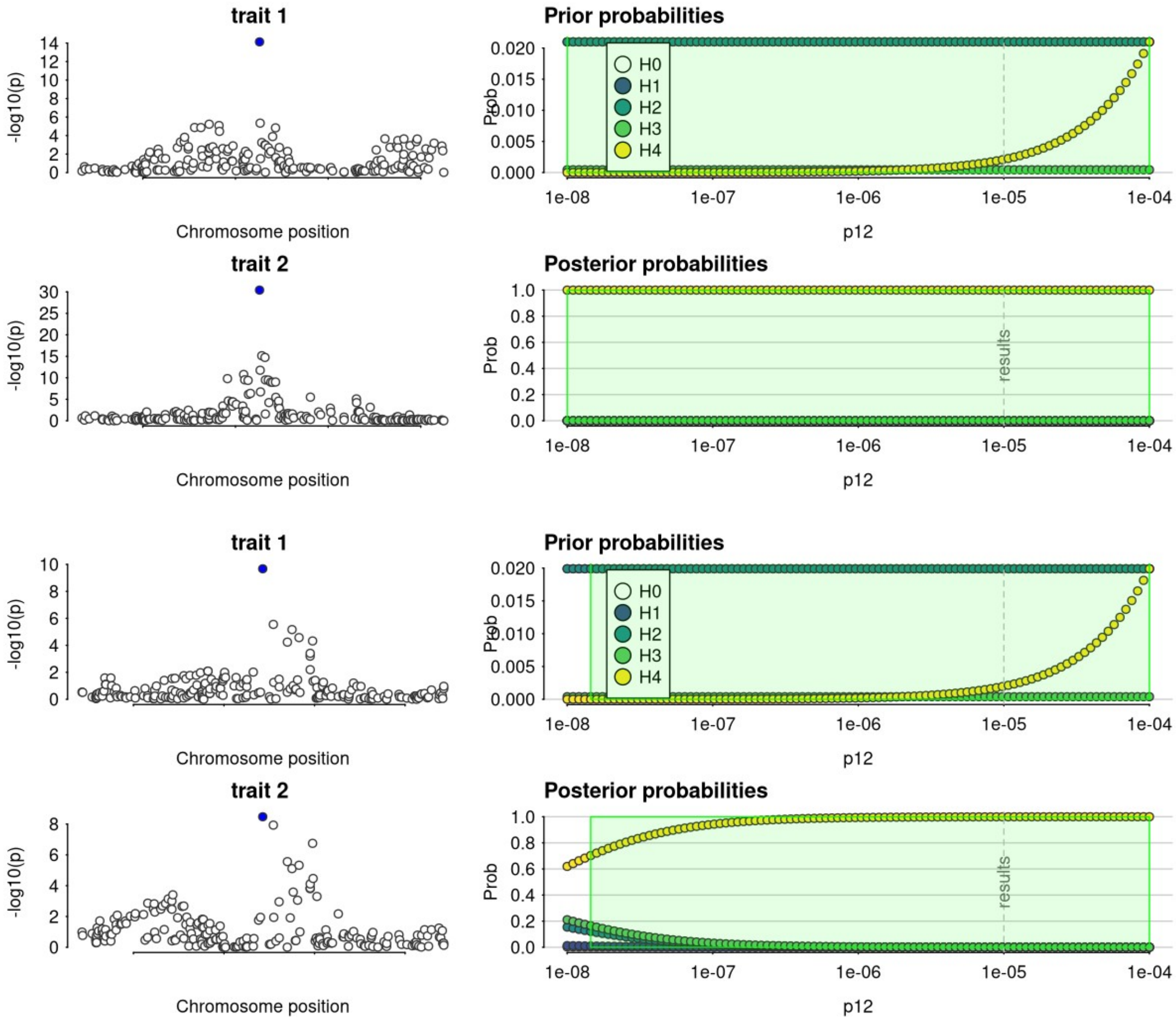


**Figure S5.** Prior sensitivity: PPH4 as a function of the shared-effect prior p12 for the DSP (AT2) and NRP2 (AT2) colocalizations (upper, DSP; lower, NRP2). At a more conservative p12 of $1\times10^{-6}$, only these two colocalizations retain PPH4 > 0.7.
